# Circulating inflammatory proteins may be potential drug targets for Idiopathic Membranous Nephropathy: proteome-wide Mendelian Randomization and colocalization analysis

**DOI:** 10.1101/2023.12.11.23299722

**Authors:** Zhihang Su, Qingqing Rao, Di Wu, Zheng Yin, Wen Liu, Qijun Wan

**Author notes:** **Correspondence:** Corresponding Author: Qijun Wan.

## Abstract

**Background:** Idiopathic membranous nephropathy (IMN) is a predominant cause of nephrotic syndrome among adults. Existing drugs are ineffective in about one-third of IMN patients, and the high recurrence rate makes them far from satisfactory. Therefore, it is imperative to find new therapeutic targets for membranous nephropathy. Circulating inflammatory proteins in plasma have been found to be related to the disease and prognosis of IMN patients, yet the causal relationship between them still remains unclear. A better understanding of the inflammatory response of IMN can help us better understand the occurrence of IMN, as well as a good way to find new therapeutic targets. In this study, we aim to use proteome-wide Mendelian Randomization and colocalization analysis to identify plasma inflammatory proteins as potential therapeutic targets for IMN.

**Methods:** We selected the genetic instrumental variables (IVs) of 91 plasma inflammatory protein quantitative trait locus (pQTL) data obtained from 14824 European population samples by Zhao JH et al. in 2023 as exposure factors. The outcome variable was obtained from the Genome-Wide Association Study (GWAS) data on IMN, which involved 2150 cases and 5829 controls across five European cohorts. To investigate the associations between inflammatory proteins and IMN risk, we conducted a two-sample bi-directional MR analysis, sensitivity analysis, Bayesian colocalization, phenotype scanning, and analysis of the Protein-Protein Interaction (PPI) network.

**Results:** The MR analysis uncovered 2 inflammatory factors associated with IMN. TNF-beta [(Tumor Necrosis Factor-beta) (IVW, OR=1.483, 95%CI=1.186-1.853, P=0.0005, P^FDR^=0.046)] was associated with an increased risk of IMN. IL-5 [(Interleukin-5) (IVW, OR=0.482, 95%CI=0.302-0.770, P=0.002, P^FDR^=0.097)] was associated with protective effects against IMN. After False Discovery Rate multiple correction and sensitivity analysis, they remain significant. None of these proteins exhibited a reverse causal relationship. Bayesian colocalization analysis provided evidence that TNF-beta share variants with IMN [posterior probability of hypothesis 4 (PPH4) = 0.88]. Utilizing the PPI network, we identified several proteins associated with the previously mentioned inflammatory proteins. Notably, TNF-beta and IL-5 were found to be linked to Nuclear Factor Kappa B Subunit 1 (NFKB1).

**Conclusions:** Our exhaustive analysis suggests a causative impact of TNF-beta and IL-5 levels on the genetically predisposed risk of IMN. These proteins hold potential as promising therapeutic targets for IMN treatment, thus necessitating further clinical investigation.

## Introduction

Idiopathic membranous nephropathy (IMN) is a condition characterized by immune-complex mediated renal disease, accounts for 23.4% of glomerular diseases, and stands as the predominant cause of nephrotic syndrome among adults(1). The defining characteristic of IMN lesions is a marked thickening of the glomerular capillary walls This thickening is attributed to the accumulation of immune complexes on the external surface of the basement membrane (2). The accumulation of these immune complexes triggers the activation of the complement pathway, which subsequently compromises the integrity of the glomerular filtration barrier. This impairment manifests clinically as non-selective proteinuria(2). Presently, the therapeutic regimen for MN comprises drugs such as cyclophosphamide, corticosteroids, rituximab, and calcineurin inhibitors(3). It is important to note that not all patients respond to treatment, with some studies reporting complete or partial response rates ranging from 57% to 81%, and about one-third of patients who respond to treatments eventually relapse(4). Therefore, it is imperative to find new therapeutic targets for membranous nephropathy. Nonetheless, despite substantial investment in human and material resources, the success rate of new drug discovery remains suboptimal. The escalated costs of research and development contribute to the inflation of healthcare expenses. Prior research has demonstrated that establishing connections between genes and corresponding phenotypes via genetic association can steer the selection of targets and indications, offering a potential solution to the prevailing challenges(5). Investigations reveal an uptick in the proportion of drug mechanisms backed by direct genetic evidence(6). Utilizing human genetic data to cherry-pick optimal targets and indications enhances the success rate of novel drug development. Human proteins, pivotal in a gamut of biological processes, emerge as the primary category of drug targets(7). In recent years, some circulating inflammatory proteins in plasma have been found to be related to the disease and prognosis of IMN patients, which has aroused people’s attention(8). For example, clinical studies have found that the levels of IL35 are increased and related to the prognosis of MN patients(9). The expression levels of IL-6 are elevated in blood and urine of PMN patients, may be a potential drug target for MN (10).And Single-cell analysis showed that the IL-17 pathway was activated in PMN renal parenchymal cells(11), serum IL-17 levels can reflect the active status of inflammation and immunity in PMN and may be a potential therapeutic target(12). In 2022, qihan et al. summarized the cytokine network of MN and confirmed the importance of inflammatory factors, proposed that in addition to BAFF, IL-35, IL-6 and IL-17A are also worthy of research as MN biomarkers and drug therapeutic targets, and emphasized the importance of updating existing treatment regimens(8). Inflammatory exposure is also thought to be closely related to the onset and progression of kidney damage(13). However, current research is insufficient to definitively establish whether these inflammatory factors are causally linked to MN or if they represent a compensatory immune response to the disease. Elucidation of these causal relationships is crucial for understanding how inflammatory factors could potentially be targeted pharmacologically.

In contrast for observational studies, mendelian randomization(MR) can avoid confounders and reverse causality, is utilized to ascertain the existence of a causal linkage between two traits, and may be a good way to determine the intrinsic causal relationship between inflammatory factors and disease at the genetic level.(14). In recent years, MR has found extensive application in drug target identification and drug repurposing(15,16). Each genetic variant correlated with the concentration of a protein is called a protein quantitative trait locus or “protein quantitative trait locus (pQTL).” Proteome-wide Mendelian randomization is a multiomic strategy for identifying the causal protein biomarkers. The advancements in high-throughput genomics and proteomics technologies have paved the way for MR analysis to identify potential therapeutic targets for a multitude of diseases, including inflammatory bowel disease, multiple sclerosis, and common cancers(17–19). Bayesian colocalization analysis is also a statistical method that uses single nucleotide polymorphisms (SNPs) derived from GWAS as genetic factors to elucidate the correlation between phenotype and disease (10). When there is an association between genetic variation variation in a genomic region and the concentration of biomarkers and IMN, the reliability of the results can be further determined using Bayesian colocalization analysis(20).When MR suggests a causal relationship between the biomarker and IMN but no colocalization, the major genetic variant may be independently associated with the trait of interest through different mechanisms rather than by influencing the concentration of the biomarker. However, until now, few pQTL data on IMN by MR studies integrating Genome-Wide Association Study (GWAS) and plasma inflammatory proteins has been reported. In this study, we aim to use proteome-wide Mendelian Randomization and colocalization analysis to identify plasma inflammatory proteins as potential therapeutic targets for IMN.

We selected the genetic instrumental variables (IV) of 91 plasma inflammatory protein pQTL data obtained in from 14824 European population samples by Zhao JH et al. in 2023 as exposure factors (16. And the outcome variable was obtained from a GWAS focused on IMN, which involving 2150 cases and 5829 controls across five European cohorts(15). The study design is shown in Figure 1. Post the screening of instrumental variables to comply with the analysis prerequisites, we executed a Mendelian Randomization (MR) analysis. Subsequently, the MR results were further corroborated using sensitivity analysis, reverse MR analysis, Steiger filtering test, Bayesian colocalization analysis, and phenotype scanning. Lastly, we constructed an interaction network among the identified proteins and other inflammatory factors proteins, as well as the interaction network between the identified proteins and the current drug targets for IMN.

**Figure 1:**
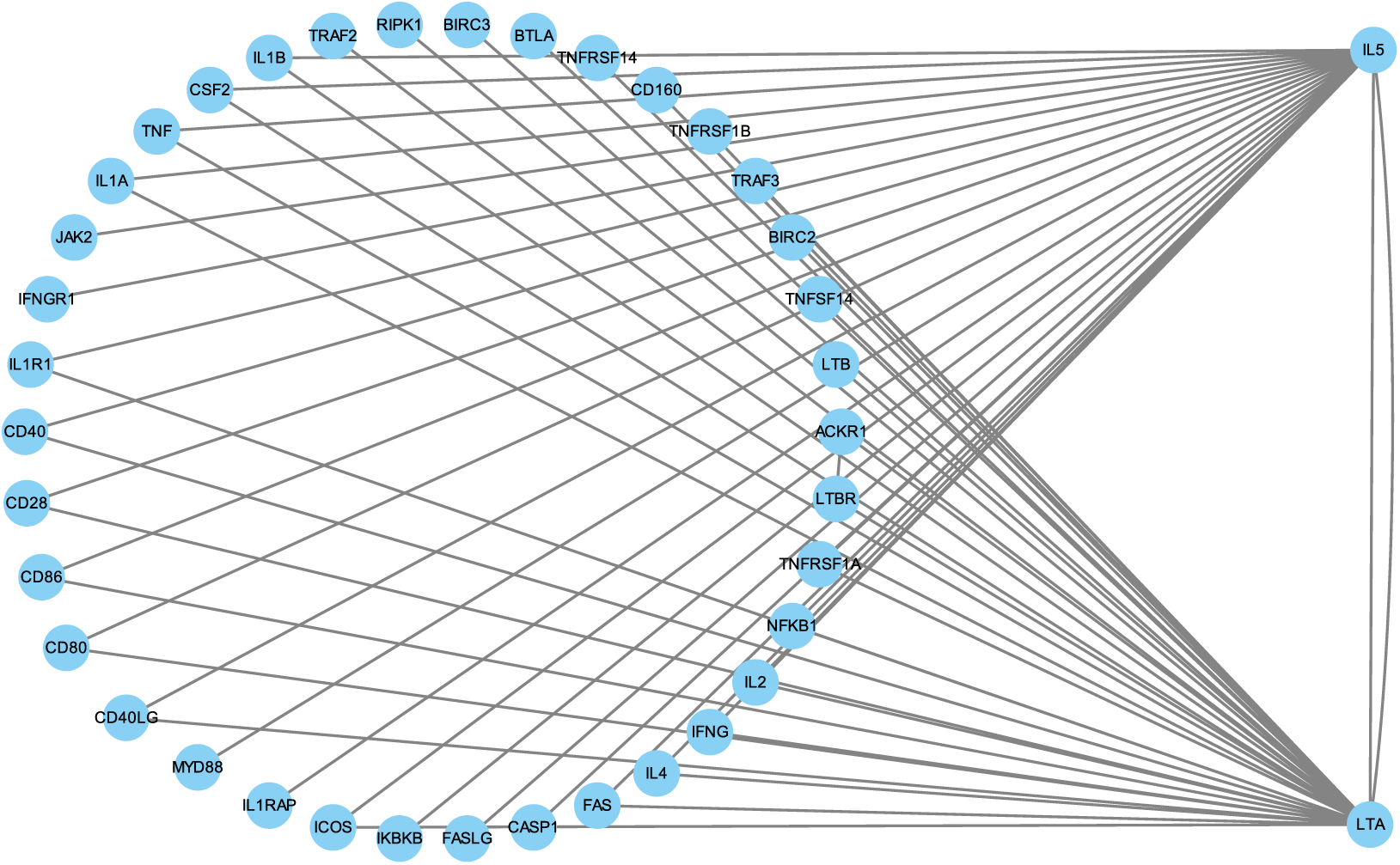
Figure 1 depicts the experimental design of this study. IVW: Inverse Variance Weighted, MR-PRESSO: Mendelian randomization pleiotropy residual sum and outlier, PPH4: posterior probability of hypothesis 4, PPI: Protein-Protein Interaction.

## Methods and Materials

### Exposure

We have selected continuous loci of inflammatory factor proteins, known as pQTLs, as the exposure variable. This data was obtained from a genome-wide protein quantitative trait locus (pQTL) study, which analyzed 91 plasma proteins using the Olink Target platform in a sample of 14,824 participants(21). The study population consisted of individuals of European descent.

### Outcome

We have chosen Idiopathic Membranous Nephropathy (IMN) as the outcome variable. The data was obtained from a Genome-Wide Association Study (GWAS) focused on primary IMN, involving 2150 cases and 5829 controls across five European cohorts(22). All patients in the database diagnosed with membranous nephropathy were confirmed through kidney biopsy, the gold standard for diagnosis. Furthermore, the database excluded secondary factors such as drugs, malignant tumors, infections, or autoimmune diseases, ensuring the focus on IMN. The study population comprised individuals of European descent.

### Selection of instrument variants

1. Relevance: It is essential to establish a strong correlation between the instrumental variables (IVs) and the inflammatory factors. We set a threshold of 5×10^-6^ for this purpose. Furthermore, we retained only those IVs that satisfied the condition F > 10 [F = R2(n - k - 1)/ k (1- R2)], ensuring a robust association between the IVs and inflammatory factors.
2. Independence: In order to ensure the independence of the variations used as IVs, we implemented a clumping process on Single Nucleotide Polymorphisms (SNPs). This was based on the Linkage Disequilibrium (LD) reference panel from the 1000 Genomes Project(23). The process involved associating any SNP within a 10000 kb window with R2 < 0.001 and retaining the SNP with the lowest p-value at each locus.
3. Exclusion: To enhance our MR analysis’s precision, we eliminated IVs that could potentially be associated with IMN (outcome p-value > 5e-05). We employed “phenoscanner” to detect confounding factors(24). Any confounder that simultaneously influenced both the exposure (inflammatory factor) and the outcome (IMN) was subsequently excluded from further analysis.

### Mendelian randomization analysis

The Inverse Variance Weighted (IVW) method, which conducts a meta-analysis of individual Wald Ratios of IVs under a multiplicative random effects model, is deemed the primary method in MR analysis. (25). Supplementary methods include MR-Egger, Weighted Median, and Weighted Mode. Results are considered meaningful when the beta values of multiple methods are consistently positive or negative, as reflected by their uniform direction on the scatter plot. Each method’s odds ratios (ORs) are quantified per standard deviation (SD) increase in the levels of risk factors(18). Essentially, our study suggests that a one SD change in the level of inflammatory factors would correspond to an OR equivalent impact on IMN’s risk or protective factors. In addressing the outcomes of the MR analysis, we will employ the False Discovery Rate (FDR) to account for multiple tests(26).

### Sensitivity analysis

MR analysis may exhibit heterogeneity and horizontal pleiotropy. To mitigate their influence on causal effects, we perform a sensitivity analysis subsequent to the MR analysis. For assessing heterogeneity, we utilize the Inverse Variance Weighted method, MR Egger, Cochran’s Q test, and Mendelian randomization pleiotropy residual sum and outlier (MR-PRESSO) to probe the internal heterogeneity of the IVs(27). If the p-value exceeds 0.05, the effect of heterogeneity on causal relationships can be disregarded. Conversely, in the presence of significant heterogeneity (p-value less than 0.05), we apply the IVW-random effects model to diminish the impact of heterogeneity on causal effects. In the case of horizontal pleiotropy, we employ the MR Egger method to ascertain its presence in the instrumental variables. Results indicating horizontal pleiotropy (p-value less than 0.05) are subsequently excluded from the analysis.

### Directional test and Reverse Mendelian Randomization analysis

We implemented a Steiger filtering test on the influence of inflammatory factors on IMN to validate the directionality of the forward MR analysis. Concurrently, a Reverse MR analysis was conducted to assess the potential impact of IMN on inflammatory factors. The roles of exposure and outcome were interchanged, with IMN serving as the exposure and inflammatory factors as the outcome for another round of reverse MR. The remaining parameters were consistent with the initial MR analysis.

### Protein-protein interaction network

We undertook a protein-protein interaction (PPI) network exploration on proteins implicated in the risk of IMN, with a nominal significance of P < 0.05 in the MR analysis. The objective was to scrutinize the interactions among prioritized proteins and ascertain if the proteins identified through MR could be associated with the target proteins of existing drugs for IMN treatment. To investigate the interactions between these significant proteins and the targets of commercially available drugs, we identified the drugs for IMN treatment and their corresponding targets from the Drugbank database based on recent reviews. All analyses were executed using the STRING tool (version 12.0) and Drugbank retrieval tools(28,29).

### Bayesian co-localization analysis

Analogous to MR, colocalization analysis serves as an alternative approach utilizing GWAS data to elucidate the relationship between traits and diseases. Despite their similarities, key differences exist. Colocalization analysis is employed to ascertain whether two phenotypes are influenced by identical or distinct causal variables. Within the scope of colocalization analysis, five hypotheses are proposed, with our primary focus being on hypotheses 3 and 4(30). Hypothesis 3 posits that the causal variable is associated with both traits, albeit with different causal variants situated within the same region. Hypothesis 4, on the other hand, suggests that the causal variable associated with both traits shares the same causal variant. A posterior probability exceeding 0.8 indicates a high degree of colocalization.

## Results

### Mendelian Randomization Results

Subsequent to MR analysis, certain inflammatory factors emerged as nominally significant. The outcome may be influenced by multiple analyses. Hence, we employed the False Discovery Rate (FDR) with thresholds set at 0.05 and 0.1 for multiple testing. While the more stringent FDR correction threshold stands at 0.05, numerous studies have incorporated results with an FDR correction threshold of 10%. At the 5% level, TNF-beta [(Tumor Necrosis Factor-beta) (IVW, OR=1.483, 95%CI=1.186-1.853, P=0.0005, P^FDR^=0.046)] was observed. At the 10% level, IL-5 (IVW, OR=0.482, 95%CI=0.302-0.770, P=0.002, P^FDR^=0.097) was noted (Figure 2 and 3). Sensitivity analysis results, which remained significant post-FDR correction, were deemed devoid of heterogeneity and horizontal pleiotropy (P>0.05).

**Figure 2:**
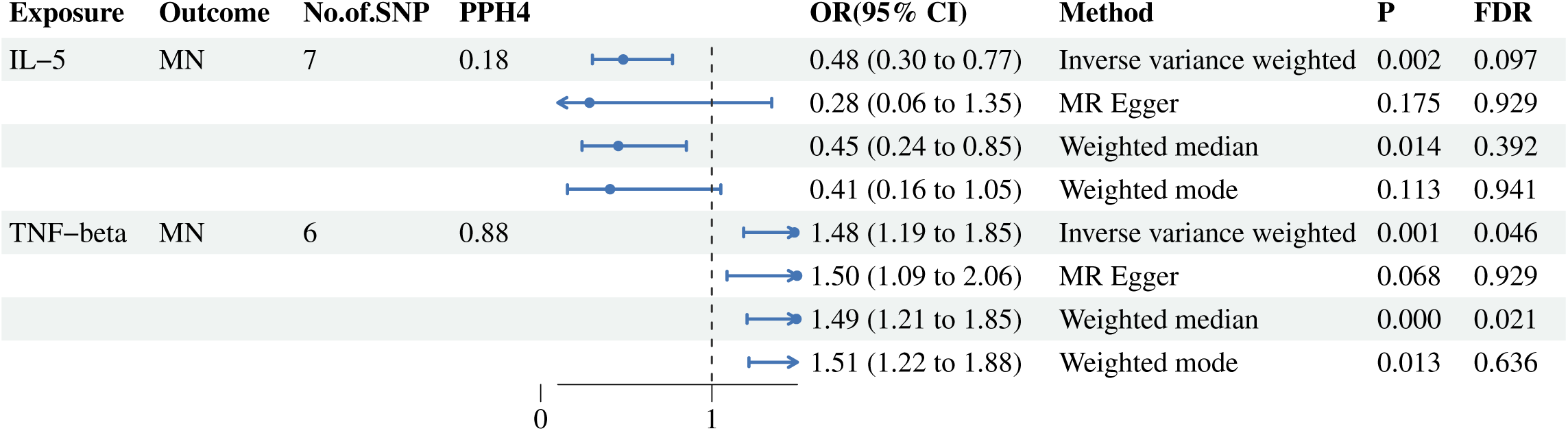
Mendelian randomization estimates of the association between inflammatory proteins and risk of membranous nephropathy. IL-5: Interleukin-5, TNF-beta: Tumor Necrosis Factor-beta, MN: Membranous nephropathy, IVW: Inverse variance weighted, OR: odds ratio, CI: confidence interval, FDR: False discovery rate, PPH4: posterior probability of hypothesis 4.

**Figure 3:**
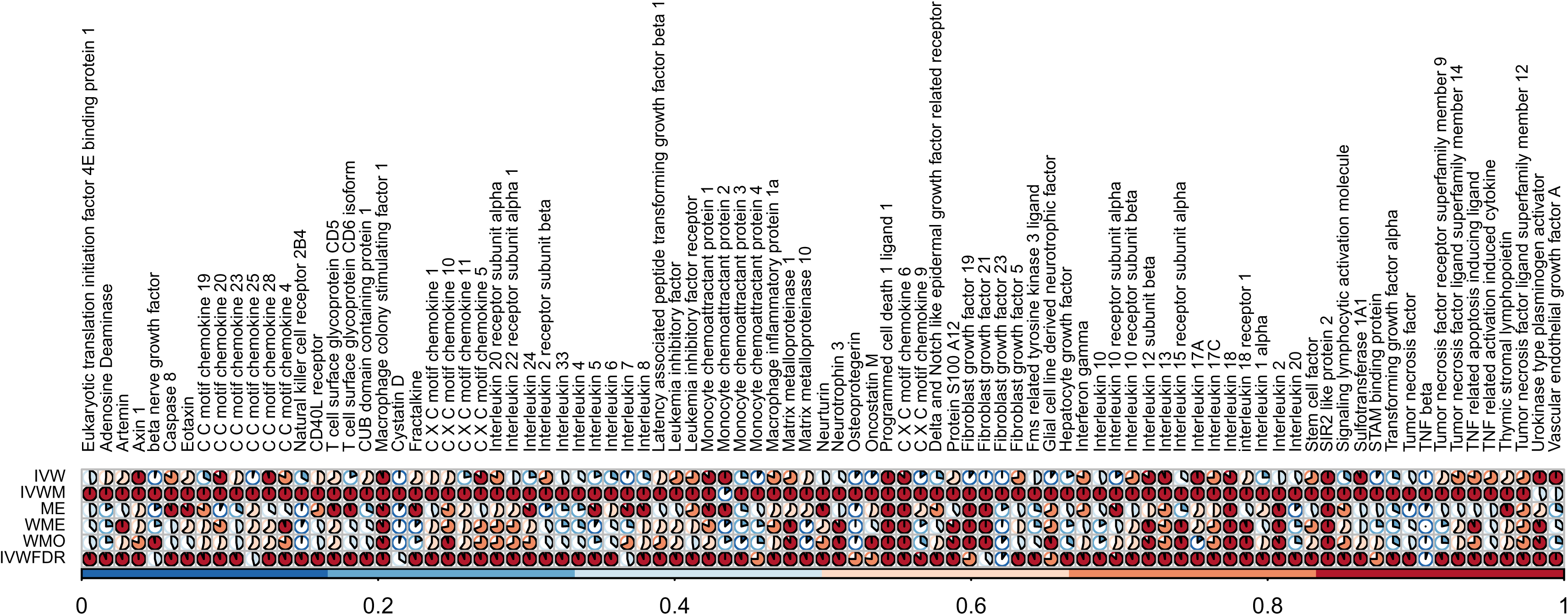
Figure 3 presents the results of all Mendelian randomization analyses conducted in this study. Some inflammatory factors lacked sufficient instrumental variables for analysis, and therefore could not be analyzed. The p-values for these factors were set to a default value of “1”. Additionally, certain inflammatory factors exhibited horizontal pleiotropy, which necessitated the use of IVWM (Inverse variance weighted with multiplicative random effects). In these cases, IVW (Inverse variance weighted) was also set to a default value of “1”. IVW: Inverse variance weighted, IVWM: Inverse variance weighted (multiplicative random effects), ME: MR-Egger, WME: Weighted median, WMO: Weighted mode, IVWFDR: Inverse variance weighted method’s p value after false discovery rate correction.

### Reverse Mendelian Randomization analysis

In the forward MR analysis, proteins deemed significant in the reverse analysis were found to be inconsequential, indicating an absence of a reverse causal effect. Furthermore, we identified a causal relationship between certain inflammatory proteins and IMN. Upon applying a FDR correction threshold of 0.05, Fractalkine (IVW, OR=1.031, 95%CI=1.012-1.050, P=0.001, PFDR=0.041), Interleukin-15 receptor subunit alpha (IVW, OR=1.033, 95%CI=1.013-1.054, P=0.001, PFDR=0.041), and Interleukin-1-alpha (IVW, OR=1.034, 95%CI=1.013-1.056, P=0.001, PFDR=0.041) emerged as significant. Sensitivity analysis results, which remained significant post-FDR correction, were deemed devoid of heterogeneity and horizontal pleiotropy (P>0.05).

### Protein-protein interaction network

Within the PPI network, we discerned associations between TNF-beta and various proteins such as LTB, IL-5, CD40, CD28, CD80, and NKFB1(Figure 4). Similarly, IL-5 exhibited connections with NFKB1, JUN, FOXP3, and STAT6. Our exploration in Drugbank revealed drugs targeting TNF-beta, including an antagonist named Etanercept. This approved drug, currently under further development, is deemed suitable for treating active rheumatoid arthritis, psoriasis, and ankylosing spondylitis. Reslizumab, Mepolizumab, and Pranlukast are identified as antagonists for IL-5. Reslizumab, an approved medication, is considered appropriate for severe eosinophilic asthma. Mepolizumab, another approved drug, is suitable for patients with severe eosinophilic asthma, eosinophilic granulomatosis with polyangiitis (EGPA), and hypereosinophilic syndrome. Pranlukast, a drug under development, is deemed suitable for allergic rhinitis (AR) and asthma.

**Figure 4:**
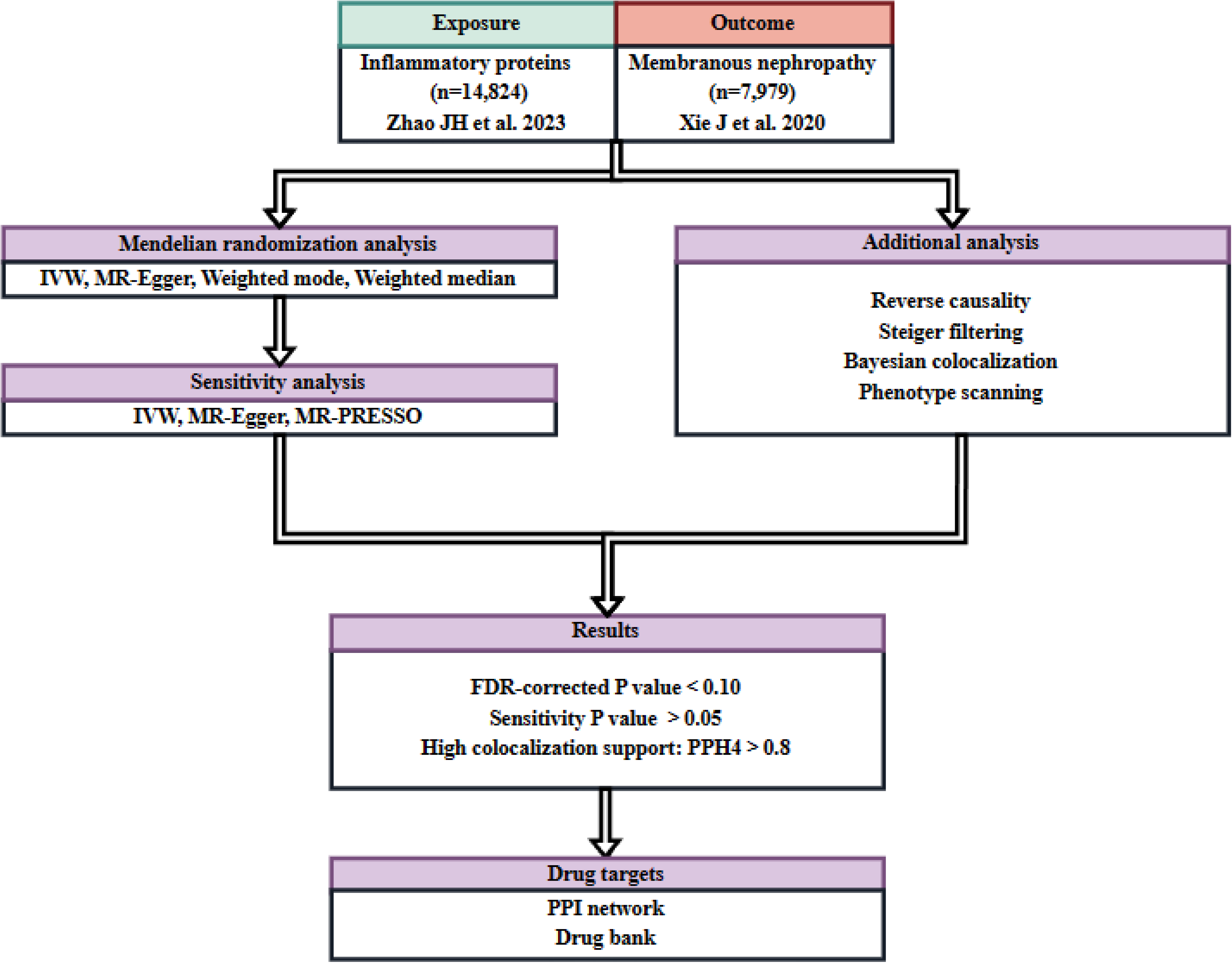
Proteins that are associated with Interleukin-5 (IL-5) and Tumor Necrosis Factor-beta (TNF-beta) in protein-protein interaction (PPI) networks.

### Bayesian co-localization analysis

Following the MR results, we performed a colocalization analysis on the significant proteins. In the case of TNF-beta, our analysis revealed a strong colocalization between rs2904602 in TNF-beta and IMN, as indicated by a Posterior Probability of Hypothesis 4 (PPH4) of 0.88. This finding suggests that they share the same causal variant. However, no meaningful colocalization was observed in the analysis of Interleukin-5 (IL-5). According to “Phenoscanner”, TNF-beta (rs2904602) is homologous to the *TBC1D22B* gene.

### Data availability

The proteome quantitative trait loci (pQTL) data for inflammatory factors was obtained from public research.(21) Data on IMN was sourced from the Kiryluk Lab.(22) We conducted our analysis using R language, version 4.3.0. The analysis involved the use of several packages in R, including “coloc” (available at https://github.com/chr1swallace/coloc.git)), “TwoSampleMR” (available at https://github.com/MRCIEU/TwoSampleMR.git), “MendelR”, and “locuscompare”.(31)

## Discussion

To better understand the inflammatory response of IMN, we conducted bidirectional Mendelian Randomization (MR) and colocalization analyses to evaluate the causal association between circulating levels of inflammatory factors and membranous nephropathy. Following FDR correction, we observed that higher genetically determined levels of TNF-β were associated with an elevated risk of membranous nephropathy, whereas IL-5 was potentially protective against membranous nephropathy. What’s more, in order to assess the potential heterogeneity and horizontal pleiotropy in MR analysis, we performed sensitivity analysis, including Inverse variance weighted, MR Egger, and Cochran’s Q test, and they still remain significant. Bidirectional MR analysis enables the differentiation of upstream and downstream components of the disease pathway, eliminating the possibility of reverse causation. Hence, bidirectional MR was conducted in the study and the TNFbeta and IL5, plasma inflammatory proteins identified by primary MR Analysis, did not show reverse causality. In this reverse MR analysis, our findings also support a causal association between membranous nephropathy and increased levels of Fractalkine, Interleukin-15 receptor subunit alpha, and Interleukin-1-alpha, implying a potential pro-inflammatory role of membranous nephropathy. In addition, Bayesian co-localization provided evidence that TNF-beta share variants with IMN [posterior probability of hypothesis 4 (PPH4) = 0.88). Ultimately, we have identified two proteins, TNF-beta and IL-5, as potential therapeutic targets for MN. PPI play a crucial role in understanding biological processes within cells. Utilizing the PPI network, we identified TNF-beta and IL-5 were found to be linked to Nuclear Factor Kappa B Subunit 1 (NFKB1).

### TNF**β** in IMN

The inflammatory cytokine tumor necrosis factor (TNF) is central to coordinating the inflammatory immune response. It plays a key role in innate immunity by inducing the expression of multiple genes involved in the inflammatory response(32). TNF can be divided into TNFα and TNFβ,both of which have similar biological potencies, which may be related to the similarity of their molecular structures and the binding of both to the same TNF receptor(33).

TNFβ also known as lymphotoxin-α, is a pleiotropic cytokine, which is primarily produced by activated T cells(34). Similar to TNF-α, TNF-β activates both classical and non-canonical NF-κB signaling by binding to TNF receptors (TNFRs)(35).NF-κB is a transcription factor that plays a crucial role in immune response, inflammation (36). and the development of various diseases. Previous studies have found TNF-β plays a crucial role in activating the NF-κB signaling pathway in immune response, contributing to the regulation of gene expression and the development of immune cells(37). Consistent with previous studies, TNF-β is associated with NFKB1 in our PPI network (Figure and Table). The activation of TNFRs has been reported that can ultimately lead to renal impairment. The circulating tumor necrosis factor receptors (cTNFRs) are associated with renal tubular TNFRs expression, and it can be a prognostic biomarkers of IMN with nephrotic syndrome(38). Inhibition of TNF signaling has been reported to attenuate renal immune cell infiltration in mice with membranous nephropathy models(39). Simultaneously, a study has revealed that TNF-beta and mRNA are connected through shared causal mutations, indicating that plasma TNF-beta (a pQTL) reflects the transcriptome—specifically, cellular secretion and tissue secretion into circulation—in various tissues. This finding aids in identifying potential genes linked to TNF-beta loci for the purpose of treating MN. Although it has been previously reported that TNFα is expressed in glomerular visceral epithelial cells in IMN patient(40), serum and urine TNFα levels are higher in IMN patients than in healthy patients(12). And a study found serum TNF-α is increase in IMN patients treated with rituximab(41). However, targeted TNFa blocking therapy has no significant therapeutic effect on reducing proteinuria(39). Qihan et al. summarized and analyzed the IMN papers related to TNF-α and think that the treatment of TNF-α may no longer be suitable for PMN(8). Considering the significant functional and structural similarity between TNFβ and TNFα, as well as the involvement of TNFR in mediating membranous nephropathy, TNFβ may be an important ligand for TNFR-mediated IMN.

To the best of our knowledge, there are no studies on the association between TNFβ and IMN. In our study, we established a link between TNFβ and an increased risk of IMN. In the colocalization analysis, the high-intensity mapping of TNFβ and IMN further confirmed the causal effect, indicating that the causal relationship between the genetic variation of TNFβ on IMN may be related by affecting the concentration of TNFβ. In addition, our PPI network further confirmed that TNFβ is related to the NFkB pathway, which has been found to be involved in the occurrence of IMN(2)., suggesting that the pathogenic effect of TNFβ on IMN may be mediated by the NFkB pathway.

Inhibiting NFκB signaling has potential therapeutic applications in cancer and inflammatory diseases. Therefore, we sopposed that inhibiting TNFβ is possible to prevent the activation of NFκB signaling pathway to treat IMN.

### IL5 in IMN

Interleukin-5 (IL-5) is a member of Th2 (type 2 inflammatory factor) cytokines, consisting of 115 amino acids, and is a disulfide-linked dimeric glycoprotein(42). IL-5 is currently considered to be one of the key drivers of the Th2 pathway and plays an important role in autoimmune diseases(42). The main physiological functions of IL5 are to promote the differentiation of B cells, activate B cells to become immunoglobulin-producing cells, and promote the proliferation, differentiation, migration, activation and survival of eosinophils(43). It has been recognized as an important target for the treatment of asthma(44). Interestingly, we found that IL-5 may be a protective factor for IMN. Through literature review, we have not found any research related to IL-5 and IMN, which is worthy of further exploration.

The causal genes of IMN have not yet been fully elucidated. When the same variations in the sequence are associated with disease risk and protein levels, the causal genes can be identified at this site. In addition, if the variations affecting protein levels also affect disease risk, then protein levels are likely to play a role in the pathogenesis of the disease rather than as a consequence(45). Conversely, in reverse analysis, if the variations affecting MN affect protein levels, then this may act as a downstream mediator of the disease. In summary, this study combines proteomics and disease in large-scale population data, using existing resources to better understand the pathogenesis of the disease, which is of great significance for the discovery of drug targets and biomarkers, and assists in drug development and re-use.

However, our study has certain limitations. The scope of this study is limited to the European population, and additional research is necessary to generalize these findings to the global human population. This study incorporates a comprehensive set of 91 inflammatory markers, representing the most recent and extensive data from existing research on inflammatory marker genome-wide association studies (GWAS). However, it should be noted that this selection does not encompass the entirety of all inflammatory markers.

## Data Availability

All data produced in the present study are available upon reasonable request to the authors.

https://gwas.mrcieu.ac.uk/datasets/ebi-a-GCST010005/

https://www.ebi.ac.uk/gwas/publications/37563310

